# Chromosome 1p deletion in colorectal cancer and lower grade glioma: possible relationship with the enteric nervous system

**DOI:** 10.1101/2023.11.07.23298214

**Authors:** Steven Lehrer, Peter H. Rheinstein

## Abstract

**Background:** Enteric neurons and enteric glial cells are a part of the enteric nervous system, which is sometimes referred to as the “second brain” of the body. This complex network of neurons controls various functions of the gastrointestinal tract, including motility, secretion, and blood flow. Research has shown that there is a connection between enteric neurons and the development of colorectal cancer, although the exact mechanisms are still being studied.

**Methods:** Because of the potential influence of chromosome mutations that may be common to both gliomas and colorectal cancer, we used the Cancer Genome Atlas (TCGA) to examine these mutations.

**Results:** 166 of 506 lower grade gliomas had the 1p 19q co-deletion. 150 of 616 colorectal cancers had a 1p deletion but no 19q deletion.

**Conclusion:** Colorectal cancer cells adhere to and migrate along the neurons of the enteric nervous system. Therefore, cancer cells might be expected to pick up mutations from neurons and enteric glial cells during recombination events. We hypothesize that the chromosome 1p deletion in colorectal cancer above is not a chance event and instead was acquired from adjacent enteric glial cells. Chromosome 1p co-deletion may confer better survival in patients with lower grade glioma in part because of loss of the MycBP oncogene, which is important in glioma development. Enteric glia might have the chromosome 1p deletion but lack the chromosome 19q deletion of CNS gliomas, making them much less vulnerable to malignant transformation than CNS gliomas. Indeed, evidence exists for a tumor suppressor gene on chromosome 19q associated with human astrocytomas, oligodendrogliomas, and mixed gliomas.

---

Eighty percent of malignant primary brain tumors are gliomas. They arise from mutations affecting neural stem cells or glial cells. One well documented pair of mutations is the chromosome 1p 19q codeletion in lower grade gliomas, a favorable prognostic marker (1).

Glial cells are present in the brain, central nervous system, and enteric nervous system, a complex network of neurons and accompanying glial cells (enteric glial cells, EGCs) which controls the major functions of the gastrointestinal (GI) tract. These cells play a crucial role in regulating intestinal motility, mucosal barrier function, and immune responses in the gut. Local glial cells may be major contributors to inflammatory pain (2) and multiple subtypes have been identified (3, 4). EGCs resemble brain glia in many ways and can function as intestinal stem cells. They are found within the walls of the entire GI tract (5-7).

Colorectal cancer is a type of cancer that originates in the colon or rectum. It is one of the most common forms of cancer and can be influenced by various genetic and environmental factors. Chromosome mutations can be a contributing factor to the development of colorectal cancer. Mutations in specific genes, such as APC (adenomatous polyposis coli), KRAS, TP53, and others, are commonly associated with colorectal cancer. In one study, deletion of chromosome 1p was detected in 22 of 82 colorectal cancers and conferred a worse prognosis (8).

Some studies have analyzed the relationship between enteric glial cells and their promotion of the development of colorectal cancer (9). Because of the potential influence of chromosome mutations that may be common to both gliomas and colorectal cancer, we used the Cancer Genome Atlas (TCGA) to examine these mutations.

## Methods

Over 20,000 primary cancer and matched normal samples from 33 different cancer types are molecularly described by The Cancer Genome Atlas (TCGA). The National Human Genome Research Institute and National Cancer Institute started working together on this project in 2006. Over 2.5 petabytes of genomic, epigenomic, transcriptomic, and proteomic data have been produced by TCGA (10). In the current study we used the TCGA colon and rectal data set (COAD) and the GDC TCGA Lower Grade Glioma (LGG) data set.

To access TCGA data we used the Xena platform (11) and cBioportal (12). Statistical analysis was done with SPSS v26.

## Results

Table 1 contains demographics and clinical characteristics of lower grade glioma and colorectal cancers studied.

**Table 1.** Demographics, clinical characteristics of lower grade glioma and colorectal cancer patients studied.

|  |  |  |  |
| --- | --- | --- | --- |
| Colorectal cancer | 616 patients | Lower grade glioma | 506 patients |
| male | 53% | male | 55% |
| female | 47% | female | 45% |
| white | 56% | white | 92% |
| other race | 44% | other race | 8% |
| American Joint Committee on Cancer Staging |  | Tumor site |  |
| T1 | 3% | Supratentorial frontal lobe | 58% |
| T2 | 17% | Supratentorial temporal lobe | 28% |
| T3 | 67% | Supratentorial parietal lobe | 9% |
| T4 | 5% | Supratentorial NOS | 2% |
| T4a | 4% |  |  |
| T4b | 1% |  |  |
| NA | 1% |  |  |
| Cancer type |  | Cancer type |  |
| colon adenocarcinoma | 63% | oligoastrocytoma | 25% |
| rectal adenocarcinoma | 26% | anaplastic astrocytoma | 24% |
| mucinous adenocarcinoma | 10% | oligodendroglioma | 23% |
| colorectal adenocarcinoma | 1% | anaplastic oligoastrocytoma | 15% |
|  |  | astrocytoma | 13% |
|  |  | diffuse glioma | 0.2% |

Figure 1 shows age at diagnosis of lower grade glioma and colorectal cancers studied.

**Figure 1.**
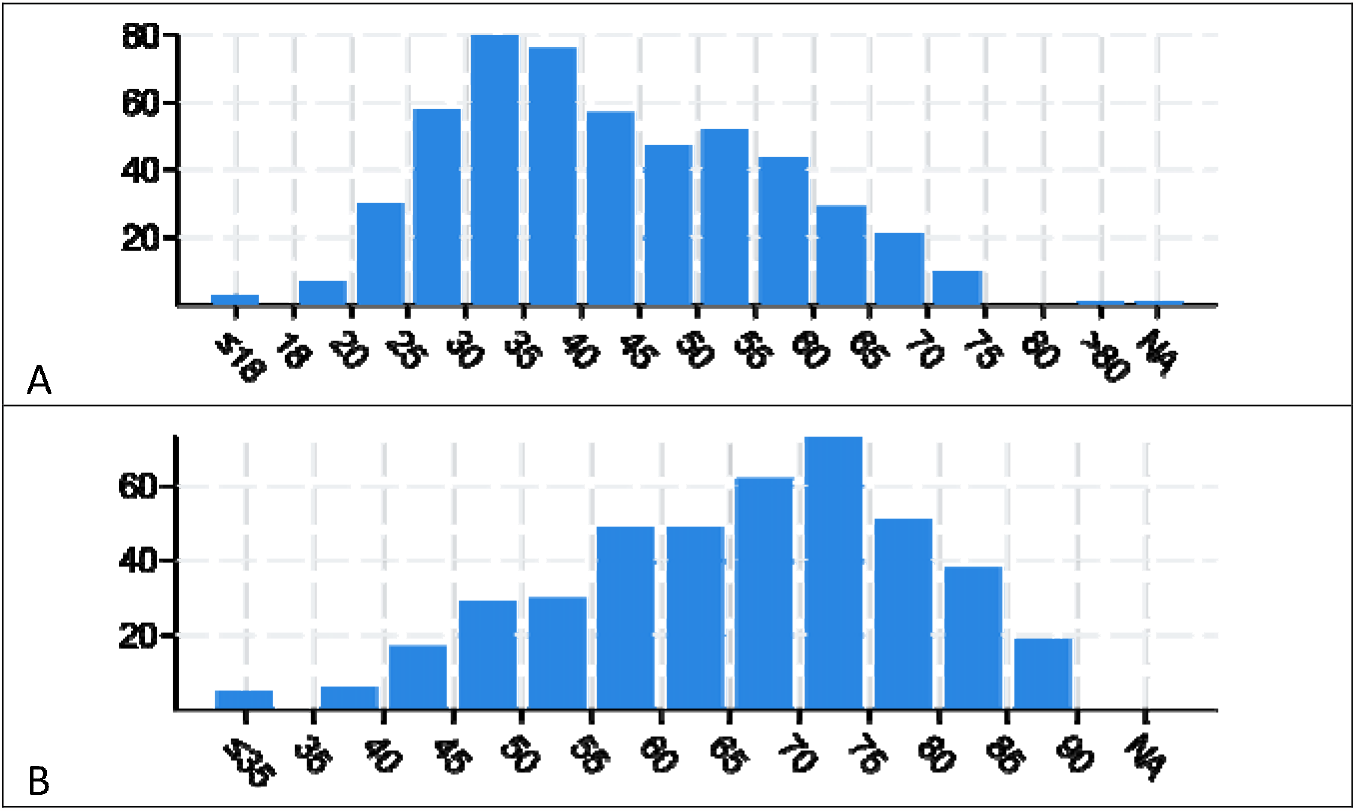
A. Age at diagnosis of lower grade glioma subjects. 1B. Age at diagnosis of colorectal cancer subjects.

Figure 2 shows overall survival and disease-free survival of lower grade glioma and colorectal cancers studied.

**Figure 2.**
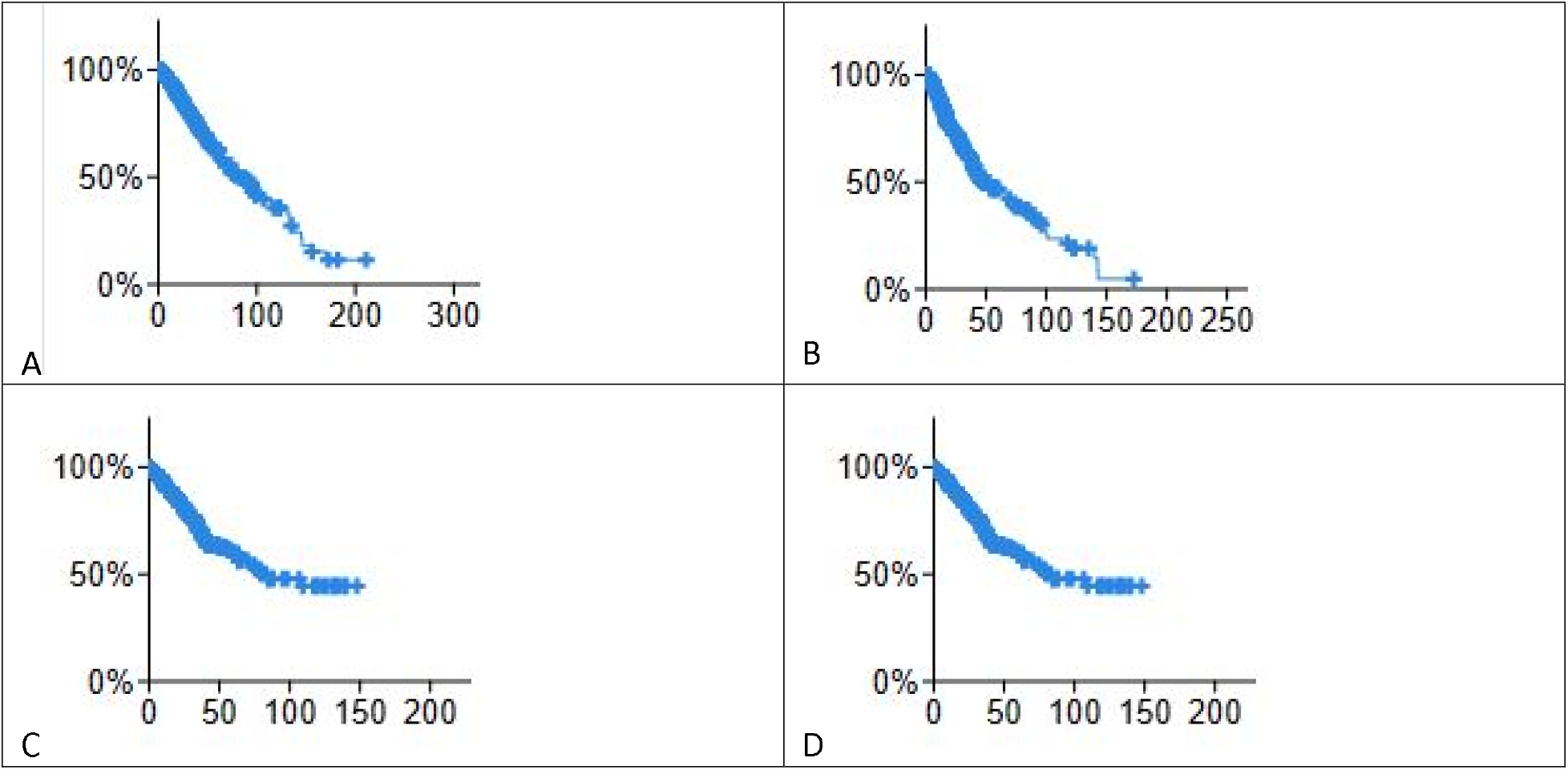
KM plots, lower grade glioma A) overall survival months B) disease free months. Colorectal cancer patients. C) overall survival months D) Disease free months.

Figure 3 shows genetic separation of lower grade gliomas into two disease groups. Each row contains data from a single sample. Row order is determined by sorting the rows by their column values. Each gray or white band in column A indicates 10 samples. Loss of chromosome arms 1p and 19q are indicated by blue blocks, columns B and C, 166 of 506 patients. TP53 and ATRX mutations are indicated in columns D and E. Anaplastic oligodendrogliomas and mixed gliomas (column F) with 1p 19q co-deletions and few or no TP53 or ATRX mutations fall into disease group 1. Anaplastic astrocytomas (orange bands column F) with no 1p 19q co-deletions and many TP53 and ATRX mutations fall into disease group 2. Although TCGA itself has played a pivotal role in developing the 2021 WHO classification (WHO CNS5 classification), its proprietary databases still retain outdated diagnoses which frequently appear incorrect and misleading according to the WHO CNS5 standards (13). It is better to classify the tumors based on 2021 WHO classification and avoid terms such as anaplastic or mixed gliomas found in TCGA.

**Figure 3.**
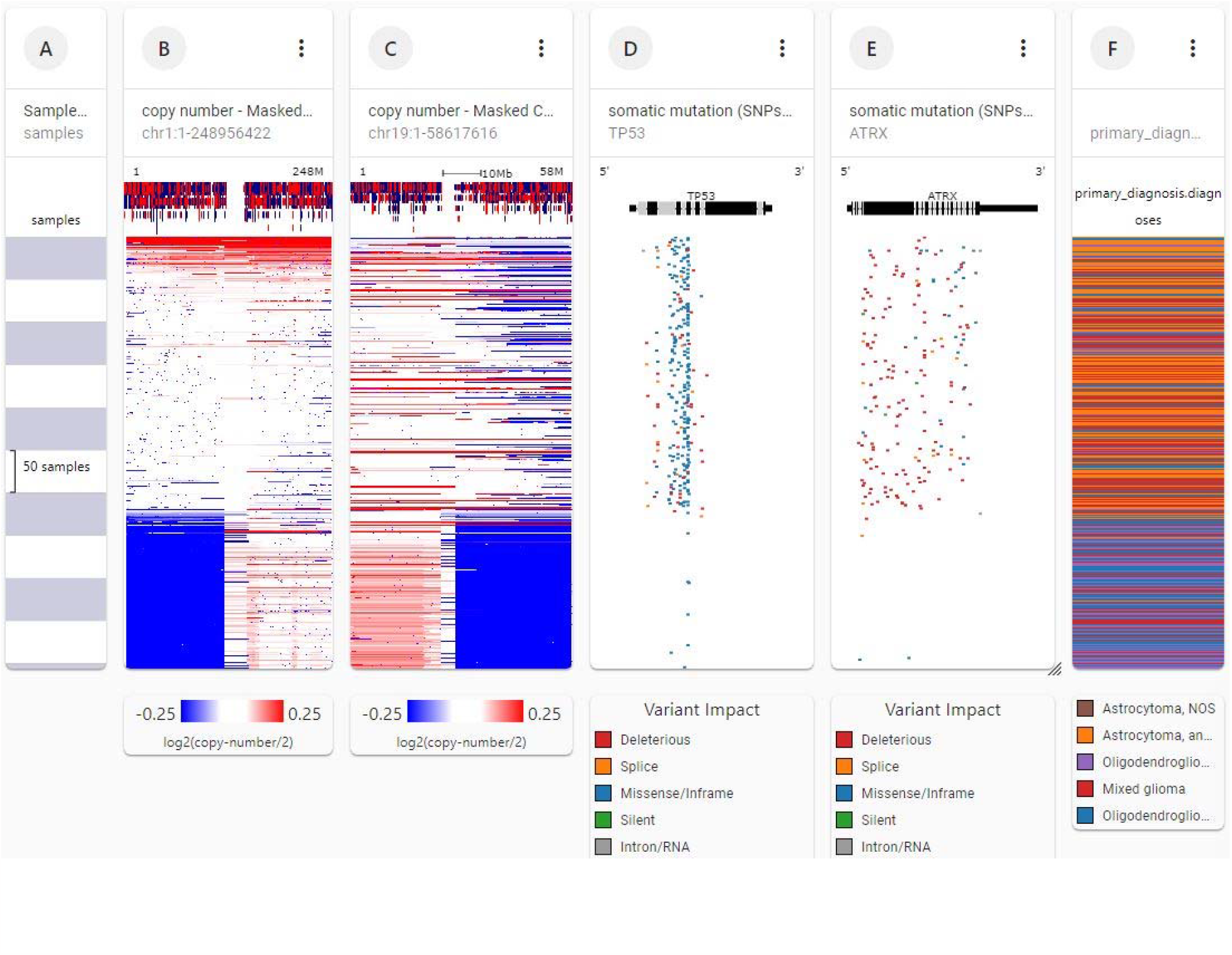
Genetic separation of lower grade gliomas into two disease groups. Each row contains data from a single sample. Row order is determined by sorting the rows by their column values. Each gray or white band in column A indicates 10 samples. Loss of chromosome arms 1p and 19q are indicated by blue blocks, columns B and C. TP53 and ATRX mutations are indicated in columns D and E, 166 of 506 patients. Anaplastic oligodendrogliomas and mixed gliomas (column F) with 1p 19q co-deletions and few or no TP53 or ATRX mutations fall into disease group 1. Anaplastic astrocytomas (orange bands column F) with no 1p 19q co-deletions and many TP53 and ATRX mutations fall into disease group 2 (UCSC Xena http://xena.ucsc.edu).

Figure 4 shows genetic analysis of chromosomes 1 and 19 in TCGA colorectal data, 616 patients. Note the loss of chromosome arm 1p (column B, lower blue block, left). The 1p loss is associated with histologic code 8140/3, adenocarcinoma not otherwise specified (column F), 150 of 616 patients. No loss of chromosome 19q, like that in lower grade gliomas, has occurred (column C). Unlike in glioma, TP53 mutations are associated with the 1p loss (column D), but ATRX mutations are not (column E).

**Figure 4.**
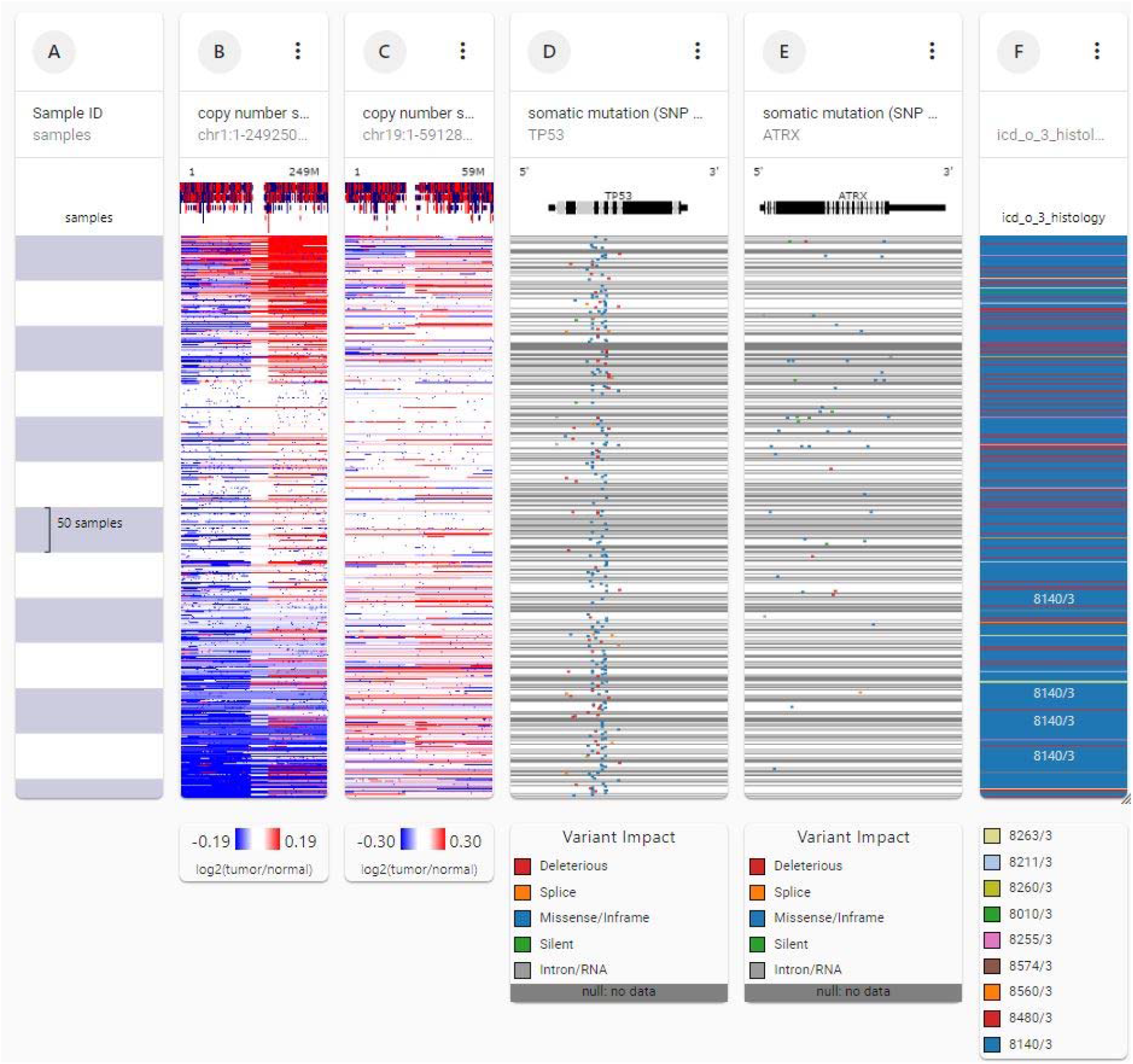
Genetic analysis of chromosomes 1 and 19 in TCGA colorectal cancer data for 616 subjects. Note the loss of chromosome 1p (column B, lower blue block, left). The 1p loss is associated with histologic code 8140/3, adenocarcinoma not otherwise specified (column F), 150 of 616 subjects. No loss of chromosome 19 has occurred (column C). Unlike glioma, TP53 mutations are associated with the 1p loss (column D), but ATRX mutations are not (column E).

Figure 5 shows survival of 616 patients with colorectal cancer. The 1p deletion had no effect on survival (p= 0.6, log rank test).

**Figure 5.**
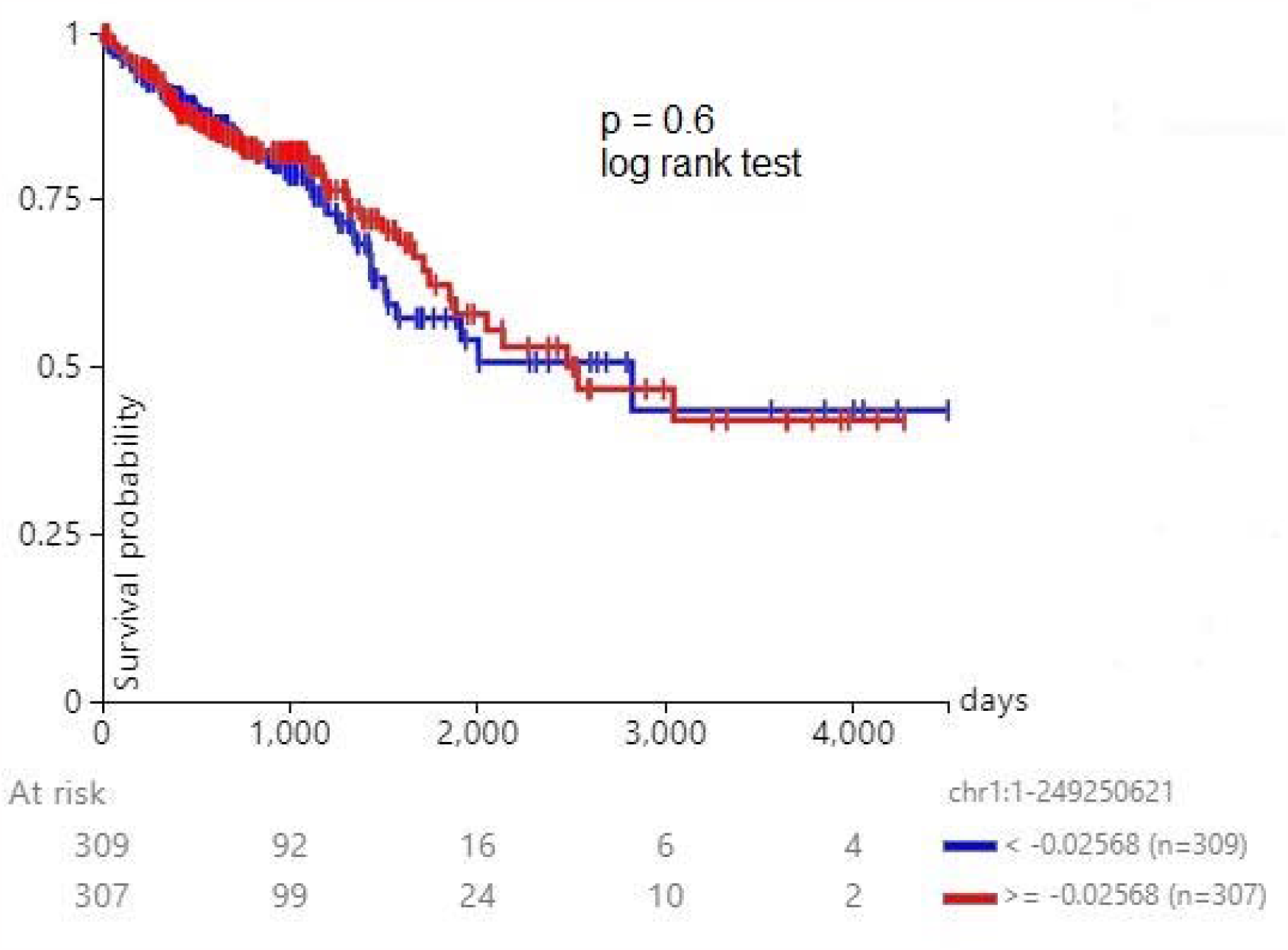
Survival of 616 patients with colorectal cancer stratified by chromosome 1p deletion. The 1p deletion had no effect on survival (p= 0.6, log rank test).

## Discussion

Enteric neurons and enteric glial cells are a part of the enteric nervous system, which is sometimes referred to as the “second brain” of the body. This complex network of neurons controls various functions of the gastrointestinal tract, including motility, secretion, and blood flow. Research has shown that there is a connection between enteric neurons and the development of colorectal cancer, although the exact mechanisms are still being studied (5).

Some potential links between enteric neurons, enteric glia and colorectal cancer include:

1. Neurotransmitter Signaling: Enteric neurons release neurotransmitters that can influence the behavior of nearby cells. Altered neurotransmitter signaling in the gut might affect the proliferation and survival of colorectal cancer cells (14).
2. Inflammation: Inflammation in the gastrointestinal tract is known to be a risk factor for colorectal cancer. Enteric neurons can influence immune responses and inflammation in the gut, potentially contributing to cancer development (15).
3. Neural Control of Motility: Abnormal motility in the colon may affect the exposure of colonic cells to carcinogens or influence the ability of the immune system to surveil and eliminate cancerous cells (16).

Colorectal cancer cells adhere to and migrate along the neurons of the enteric nervous system (17). Therefore, cancer cells might be expected to pick up mutations from neurons and enteric glial cells during recombination events. We hypothesize that the chromosome 1p deletion in colorectal cancer above is not a chance event and instead was acquired from adjacent enteric glial cells.

A persistent question is why the incidence of glioma of the enteric nervous system is so low. One possible explanation is that enteric glia might have the chromosome 1p deletion and lack the chromosome 19q deletion of CNS gliomas.

The chromosome 1p 19q co-deletion is a favorable prognostic factor in patients with low grade glioma. Chromosome 1p co-deletion may confer better survival in patients with lower grade glioma in part because of loss of the MycBP oncogene, which is important in glioma development (18).

Evidence exists for a tumor suppressor gene on chromosome 19q associated with astrocytomas, oligodendrogliomas, and mixed gliomas (19, 20). Lower grade gliomas (figure 3) had the 1p 19q codeletion or no deletions. None had only the 1p deletion, suggesting that if the 1p deletion occurs alone it may render glial cells more resistant to malignant transformation.

Our study has weaknesses:

1. chromosome 1p has a huge number of genes. An entire chromosomal arm deletion, in addition to the very different embryology and epidemiology of colorectal cancer versus lower grade glioma, could result in a significant chance of false discovery of this nonspecific mutation.
2. 1p and 19q co-deletion in brain tumors is mediated by an unbalanced t(1;19)(q10;p10) chromosomal translocation (21, 22). It is a centric fusion between chromosomes 1 and 19 with subsequent loss of 1p/19q whereas the 1q/19p chromosome is retained. 1p deletion in colon cancer is different. It is of various sizes with a minimum common deleted region of 1p36 (23-26). One group found that the deleted region is between markers D1S199 and D1S234 (26) whereas another group between markers D1S2647 and D1S2644 (27). Thus, the two genetic events, 1p and 19q co-deletion in brain tumors and 1p deletion in colon cancer may not be the same and may not be related.
3. Enteric glial cells were shown to stimulate expansion of colon cancer stem cells and ability to give rise to tumors via paracrine signaling (5, 28).

In conclusion, we hypothesize that the chromosome 1p deletion in colorectal cancer is not a chance event and instead is acquired from adjacent enteric glial cells. Moreover, enteric glia might have the chromosome 1p deletion but lack the chromosome 19q deletion of CNS gliomas, making them much less vulnerable to malignant transformation than CNS gliomas.

## Data Availability

Data sources described in the article are publicly available.

https://xenabrowser.net/

